# Progressive worsening of the respiratory and gut microbiome in children during the first two months of COVID-19

**DOI:** 10.1101/2020.07.13.20152181

**Authors:** Rong Xu, Pengcheng Liu, Tao Zhang, Qunfu Wu, Mei Zeng, Yingying Ma, Xia Jin, Jin Xu, Zhigang Zhang, Chiyu Zhang

## Abstract

Children are less susceptible to COVID-19 and manifests lower morbidity and mortality after infection, for which a multitude of mechanisms may be proposed. Whether the normal development of gut-airway microbiome is affected by COVID-19 has not been evaluated. We demonstrate that COVID-19 alters the respiratory and gut microbiome of children. Alteration of the microbiome was divergent between the respiratory tract and gut, albeit the dysbiosis was dominated by genus Pseudomonas and sustained for up to 25-58 days in different individuals. The respiratory microbiome distortion persisted in 7/8 children for at least 19-24 days after discharge from the hospital. The gut microbiota showed early dysbiosis towards later restoration in some children, but not others. Disturbed development of both gut and respiratory microbiomes, and prolonged respiratory dysbiosis in children imply possible long-term complications after clinical recovery from COVID-19, such as predisposition to an increased health risk in the post-COVID-19 era.

## Introduction

COVID-19 caused by SARS-CoV-2 has impacted millions of peoples in more than 200 countries around the world^1,2^. Compared with adults, children appear to be less susceptible to COVID-19 with extremely low morbidity and mortality^3-5^; and children with COVID-19 often have mild symptoms with a faster recovery, and better prognosis. The reasons for these are not entirely clear. Early-life development and maturation of human microbiomes shape health status in later life^6-9^, and delayed development or dysbiosis of the microbiomes during childhood has been linked to predisposition to various diseases in adulthood^7-12^. The effect of COVID-19 on the gut microbiome had just began to be evaluated in adults^13,14^, but never in children. We recently evaluated the longitudinal effects of COVID-19 on both the respiratory and gut microbiome in adults, and revealed that the respiratory and gut microbiome presented a contemporaneous change from early dysbiosis towards late incomplete restoration during the course of disease (Xu et al., unpublished observation). How COVID-19 impacts on the respiratory and gut microbiomes of children is not known. Here, we report temporal dynamics of respiratory and gut microbiome in children with COVID-19.

## Results

### Study cohort

Nine COVID-19 children between 7-139 months old were enrolled in this study together with 14 age-matched healthy control children. A total of 103 specimens including 27 sets of paired specimens (at least two of throat swab, nasal swab and feces) were collected from children with COVID-19 (Supplementary Fig. 1). The children were being followed between 25-58 days after symptom onset. All samples were subjected to high-throughput sequencing of the V4-region of bacterial 16S rRNA gene (Methods).

### Respiratory and gut microbiome dynamics in COVID-19

We analyzed the 16S-rRNA gene sequences of all specimens from three body sites, and obtained 2,187 sub-OTUs (sOTUs) that represent 15 known phyla including 200 known genera (Supplementary Table 1). Using the DMM method (Supplementary method), we identified 8 community types (Fig.1a). The specimens from healthy children clustered into two community types, one bears the signature of stool samples (H-GUT), and another represents the collection of all three kinds of samples (throat swab, nasal swab and feces) (H-MIX). H-GUT is significantly separated from H-MIX (Fig. 1b), with significantly lower richness and evenness (Fig.1c). H-GUT was featured by *Moraxella*, a commensal in nasal passages of infants, implying that it might not represent a normal gut microbiome status. Because the development of infant microbiome is influenced by maternal materials from multiple sites (stool, vaginal, oral and skin) among which bacteria from maternal gut being the most important contributor^15^, infants and children may share the same or very similar microbial community structures of the nasal cavity, throat and gut^16,17^. Therefore, H-MIX represents the gut and respiratory tract microbiomes of healthy children. In fact, the nasal cavity, throat and gut still maintain similar microbial structures in adulthood.

**Fig 1.**
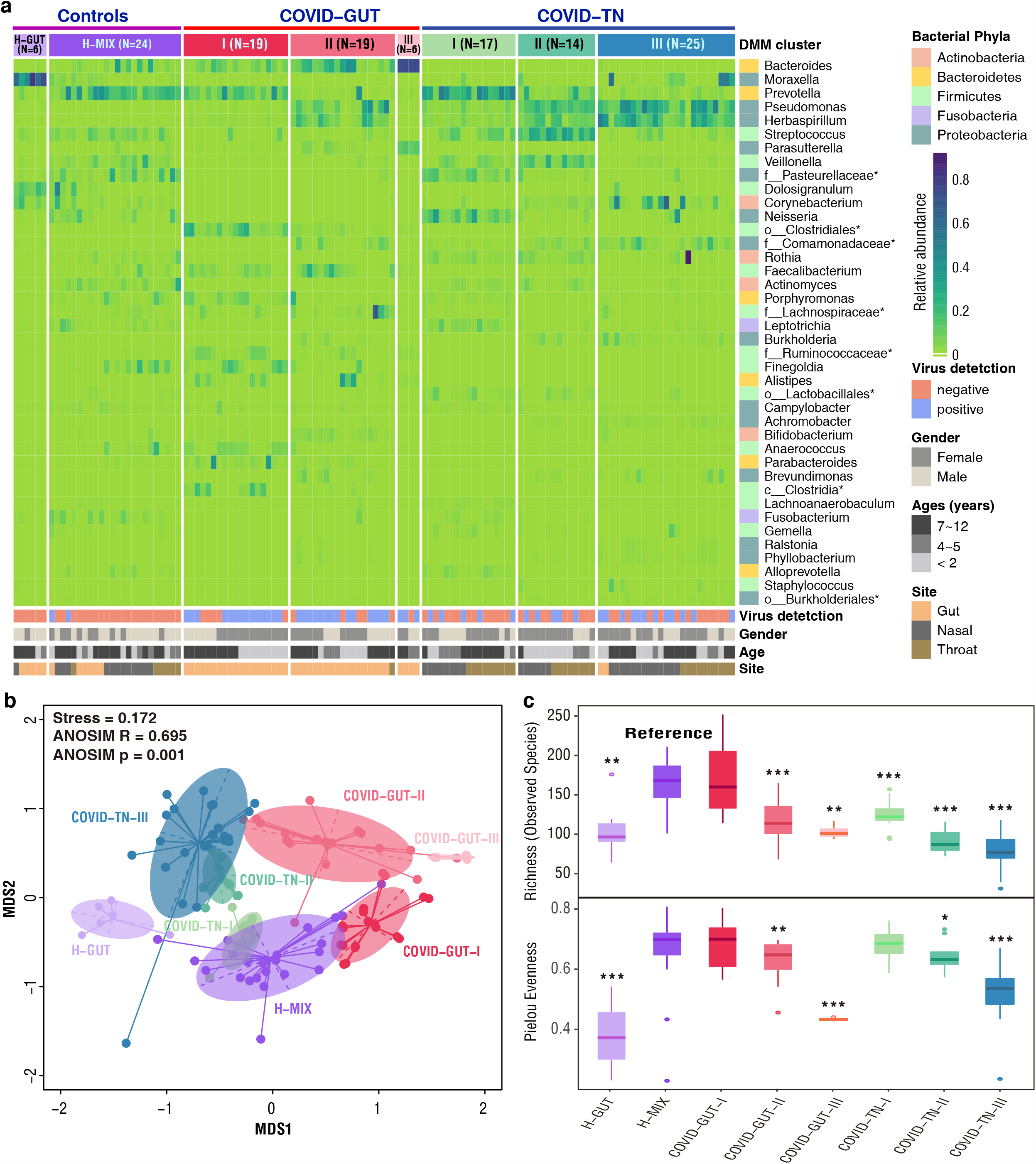
Dirichlet multinomial mixtures (DMM) clustering analyses of 16S rRNA data separate COVID-19 children into groups with distinctive features (*N* = 128). a. Eight distinct clusters were identified based on lowest Laplace approximation for control and patient samples from gut or nasal and throat cavities. Heat map showing the relative abundance of 40 most dominant bacterial genera per DMM cluster. The stars represent unclassified genera. H-GUT indicates abnormal gut microbial community structure of healthy children. H-MIX represents normal microbial community structure of a mixture of fecal, nasal and throat samples of healthy individuals. COVID-GUT I-III enriched in fecal samples of COVID-19 children; COVID-TN I-III enriched in both nasal and throat samples of COVID-19 children. b. Nonmetric multidimensional scaling (NMDS) visualization of DMM clusters using Bray-Curtis distance of bacterial genera. Significant separation of microbial community structures was implicated by the ANOSIM statistic R closer to 1 with < 0.05 P value. The stress value lower than 0.2 provides a good representation in reduced dimensions. **c**. Box plots showing the alpha diversity (richness and evenness) per each DMM cluster. Wilcoxon test was used to compare difference of H-MIX cluster and others under three significance levels of * P < 0.05, **P<0.01, and***P<0.001.

Bacteria from stool specimens of COVID-19 children fell into three distinct community types (COVID-GUT I-III), and those from nasal and throat swabs formed another three distinct types (COVID-TN I-III) (Fig. 1a). All COVID-19-related types are significantly separated from the type H-MIX except COVID-TN-I that overlaps with H-MIX (Fig. 1b). In particular, three respiratory tract-related types and three GUT-related types of COVID-19 children are also significantly separated from each other, and distinctly different from healthy children. These results indicated that SARS-CoV-2 infection significantly changed the gut and respiratory tract microbiota of children, and the separation of bacterial community structures between the gut and respiratory tracts suggested that the normal development of the microbiota may be impaired.

All COVID-19-related types showed lower richness and evenness than H-MIX, except for COVID-GUT-I that has the most similarity to H-MIX and relatively normal microbiome structure. There was a gradual decrease from community type I to III for both gut and respiratory tract (Fig. 1c), indicating a progressive deterioration (dysbiosis) of the microbiome. Overall, the dysbiosis appeared to be more severe in the respiratory tract than in the gut.

### Indicator genera of eight DMM clusters

To characterize eight microbial community types, we identified 35 indicator genera (Fig. 2a). The H-MIX type was characterized by 11 genera, and the predominant commensal bacteria contained *Prevotella, Streptococcus*, unclassified *Pasteurellaceae*, and *Actinomyces* (Fig. 2b). Some of the indicator bacteria in H-MIX were shared by the community types COVID-GUT-I (e.g. *Prevotella, Porphyromonas, Finegoldia, Anaerococcus, etc*.) and COVID-TN-I (e.g. *Prevotella, Neisseria, Fusobacterium*, unclassified *Pasteurellaceae, Leptotrichia* etc.). As a dysbiosis status, community type COVID-GUT-III was dominated by *Bacteroides*, as well as *Parasutterella* that is associated with irritable bowel syndrome and other intestinal chronic inflammation^18^. Community type COVID-TN-III was dominated by highly abundant *Pseudomonas* and *Herbaspirillum*, and it had higher levels of genera of *Corynebacterium, Comamonadaceae, Burkholderia, Achromobacter, Brevundimonas, Ralstonia, Phyllobacterium*, and *Burkholderiales* than other community types (Fig. 2b). Genus *Pseudomonas* is a notorious human pathogen associated with various diseases (e.g. pneumonia), and the samples were overwhelmed by the dominant species *Pseudomonas veronii* (100% sequence identity)^19^. Apart from COVID-TN-III, genus *Pseudomonas* also dominated community type COVID-TN-II with *Streptococcus*, and COVID-GUT-II with *Bacteroides*. Furthermore, *Achromobacter* and *Burkholderia* are associated with cystic fibrosis^20,21^, and most other genera are environmental bacteria. The predominance of *Pseudomonas* together with the colonization of various environmental bacteria in type COVID-TN-III imply an extreme dysbiosis in upper respiratory tract.

**Fig 2.**
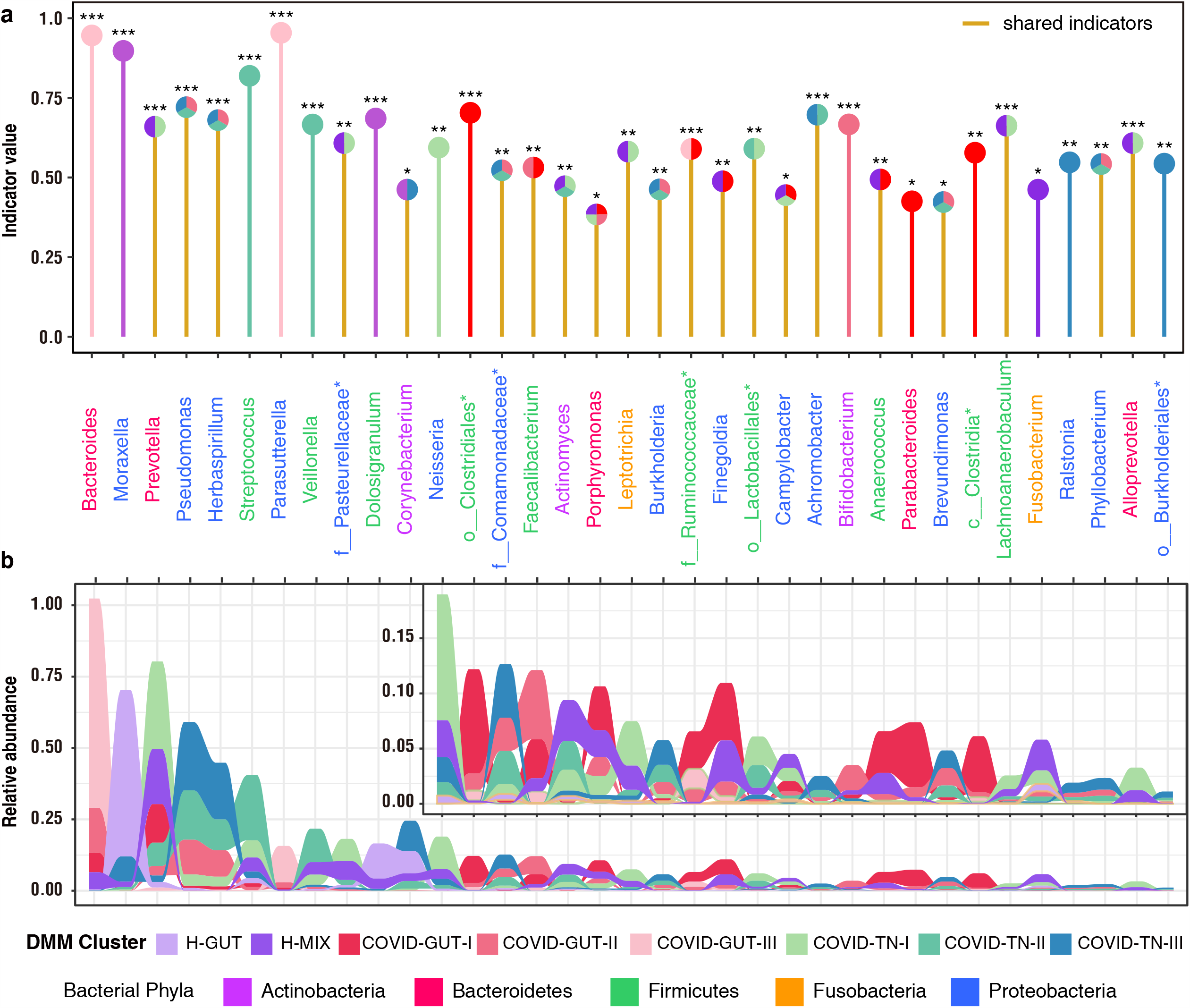
Identification (a) and relative abundance distribution (b) of indicator genera of eight DMM clusters. Indicators of eight microbial community types (DMM clusters) were identified from top 40 genus contributing to DMM clustering in Fig.1a (See Supplementary Methods). Twenty-one indicator genera are shared by two or more community types with similar diversity levels (e.g. II-III, or I-H-MIX). Significance levels of indicators are as follows: * P <0.05, ** P <0.01, and *** P < 0.001.

### The dynamic change of children during COVID-19

Recently, we observed synchronous restoration of the microbiomes of both respiratory tract and the gut towards normal structure in COVID-19 adults within a short time (6-17 days) after symptom onset (Xu et al., unpublished observation). Distinct from adults, the microbiome community compositions were extremely dynamic in children during COVID-19, and the changes of the community types in the respiratory tract and gut were divergent (Fig. 3a). The respiratory (especially nasopharyngeal) microbiome of 7/8 children (except CV05) appeared to evolve from early healthy (H-MIX) or high-diversity community structures (COVID-TN-I) to late low-diversity dysbiosis structure (COVID-TN-III), indicating a steady deterioration in composition and function of the respiratory microbiome despite a fast clinical recovery (Fig. 3a). Surprisingly, the respiratory dysbiosis was sustained at least 19-24 days after discharge (i.e., 42-58 days after symptom onset) in three children (CV01, CV02 and CV09).

**Fig 3.**
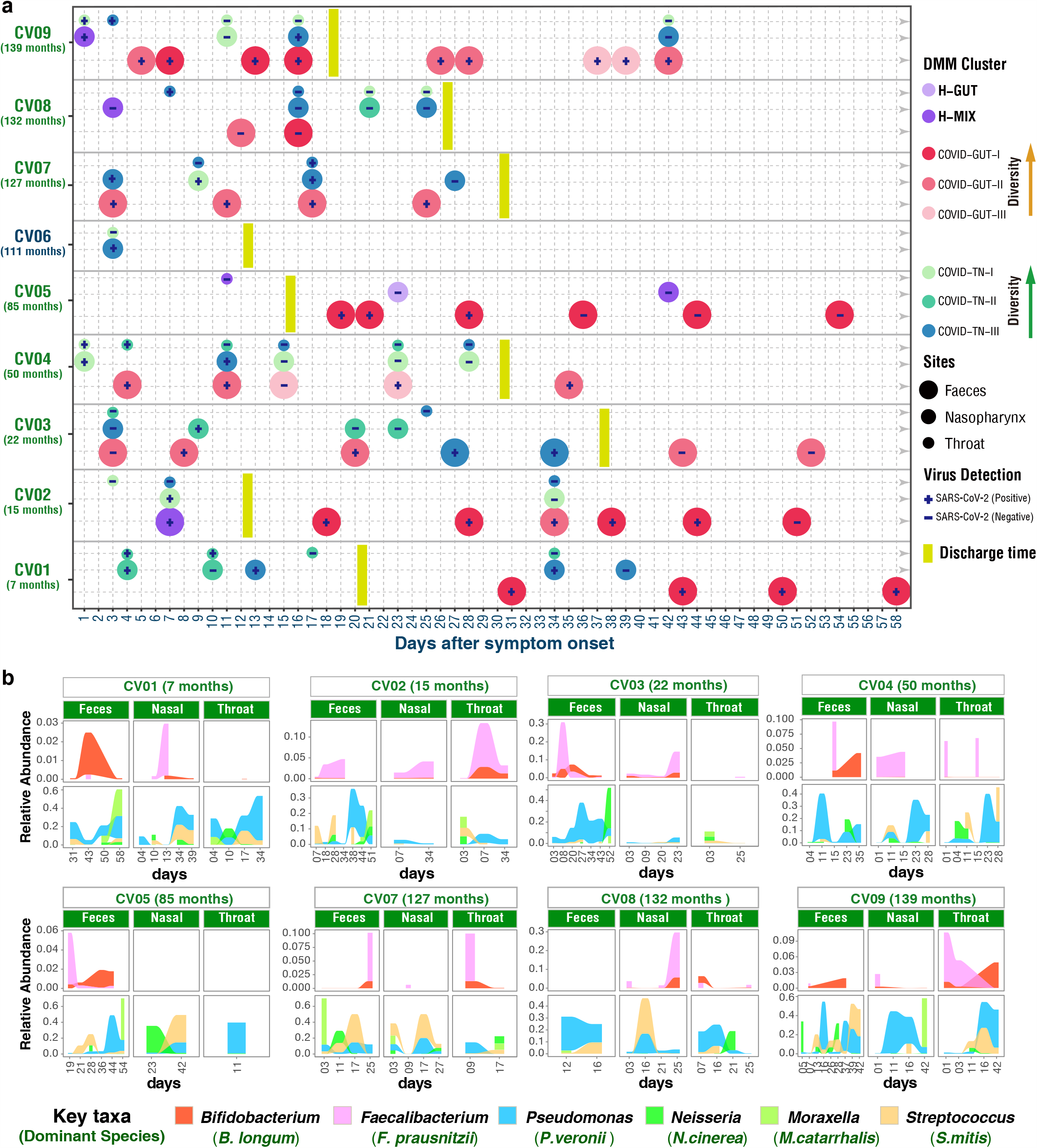
Dynamic change of both DMM clusters (a) and six key taxa (b) in three body sites of COVID-19 children. Age (months) of each COVID-19 child was shown in brackets. The community type levels were divergent between respiratory tract and gut (e.g. on days 11, 15 and 23 in CV04, and days 16 and 42 in CV09). The respiratory microbiome appeared to be progressively worsening in seven children (CV01-CV04 and CV07-CV09) for 25-42 days after symptom onset. The worsening of the gut microbiome appeared in four children (CV03, CV04, CV07 and CV09), and sustained for 25-52 days. **b**, key taxa of DMM clusters were shown in eight COVID-19 children with at least two time points of sampling. Linked to Supplementary Fig.2.

In contrast, the gut microbiome alternation varied greatly among these COVID-19 children. Improvement or restoration was observed in three children (CV01, CV02 and CV05), but a worsening trend of unstable bacterial genera occurred in another three children (CV03, CV04 and CV09) (Fig. 3a). For example, the community type of CV09 improved from COVID-GUT-II to COVID-GUT-I on day 7 after symptom onset, but deteriorated to COVID-GUT-III on day 37. For CV03, whose microbiome got worse from a gut community type COVID-GUT-II on day 19 to a respiratory community type COVID-TN-III on day 27, and returned to COVID-GUT-II on day 43. The shift from a slightly dysbiosis gut community type to a severely dysbiosis respiratory community type implies microbial translocations from respiratory tract to gut. The restoration or worsening of the gut microbiome showed no association with clinical recovery (discharge from the hospital) or the presence or absence of SARS-CoV-2 RNA in the gut (Fig.3a).

Our data clearly demonstrate a progressively worsening of microbiome in both the respiratory tract and gut of children during the course of COVID-19. The worsening was predominantly driven by *Pseudomonas* species *P*.*veronii* (Fig. 3b and Supplementary Fig. 2), which was the most prevalent *Pseudomonas* species identified, and had a relative abundance of over 20% in most COVID-19 children. Genus *Streptococcus* (mainly *S*.*mitis*) also contributed to the worsening of the microbiome^22^. On the other hand, the presence of probiotic *Bifidobacterium* and the most important butyrate-producing bacteria *Faecalibacterium* were inversely correlated with the existence of *Pseudomonas* (Fig. 3b and Supplementary Fig. 2), despite these beneficial bacteria presented at a very low relative abundance and often decreased in late disease stage.

### Bacteria–bacteria co-occurrence networks

The co-occurrence network analysis revealed significant microbial cross-talk among different body sites of children with COVID-19 (Fig. 4). There were three main co-occurrence networks identified. Positive co-occurrence relationships were observed within and between bacteria from the respiratory tract and the gut (FDR-adjusted P<0.001, Pearson correlation r > 0.4), indicating the presence of frequent bacterial cross-talk between different body sites. Similar to our observation in adults, the co-occurrence networks were relatively separated by different diversity-levels of community types, but not by body sites. For example, bacteria from the community type COVID-TN-III are closely associated with those from COVID-TN-II and COVID-GUT-II (Fig. 4). In this network, *Pseudomonas* was positively correlated with some environmental bacteria. In another two networks, probiotic (e.g. *Bifidobacterium* and *Faecalibacterium*) were mainly correlated with commensals and community types H-MIX, COVID-GUT-I, COVID-TN-I and COVID-TN-II (Fig. 4). Therefore, the community types are more representative of microbiome status.

**Fig 4.**
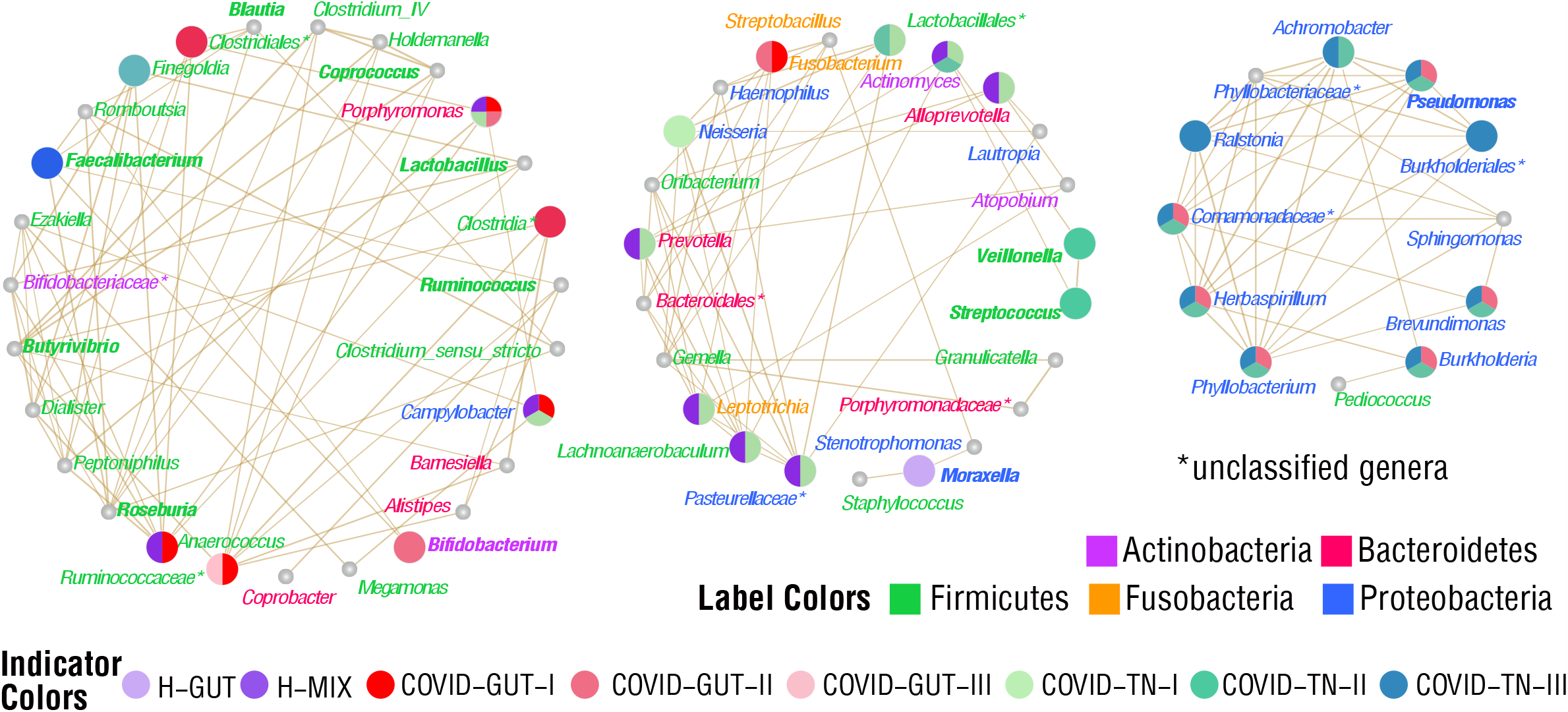
Co-occurrence network of gut, nasal and throat microbiotas in the progression of COVID-19 children. Positively correlated microbial pairs with both Pearson correlation r values higher than 0.4 and FDR-adjusted P values lower than 0.001 were shown.

## Discussion

SARS-CoV-2 infection causes damage to multiple organs either through direct infection, or indirectly disrupt host homeostasis, including perturbation of gut and respiratory microbiota in adults. Here, we report the first longitudinal investigation of microbiome with sampling from multiple anatomical sites of children with COVID-19, and present evidence that children have distinct differences compared to adults (Xu, unpublished observations), with respect to the dynamic changes of microbiota during the course of disease. The study of children is a significant feature because early-life microbiome plays important roles in the development of host immunity, metabolism, and neural systems, affecting profoundly health status in later life^7^. The microbiome attains a relatively stable adult-like structure at the age 3 after a highly dynamic initial developmental (months 3-14) phase, a transitional (months 15-30) phase, and finally a stable phase (months 31-46)^16^. The development of infants’ microbiomes is more easily influenced by various internal (genetic) and external factors (e.g. birth mode, feeding type, siblings, antibiotics, and infection)^7,9^, and maternal gut strains provide the largest contribution to the microbiome composition^15^. Early-life microbiome disruption caused by antibiotics and malnutrition is associated with increased risk of health problems later in childhood and even adulthood, such as developmental retardation, allergic diseases, obesity, diabetes and immune dysfunction^3-5,23,24^.

SARS-CoV-2 infection impaired the respiratory and gut microbiota in both adults and children, but the respiratory and gut microbiome of children and adults faced diverging fates (Supplementary Fig. 3). In some mild adult cases, the respiratory and gut microbiomes showed a synchronous restoration from early dysbiosis towards later near-normal structure within a short period (6-17 days), substantially earlier than their clinical recovery (Xu et al. unpublished observation). In children, however, the dynamic changes of the microbiome were divergent between respiratory tract and gut, and the microbiome appeared to be progressively worsening, especially in the respiratory tract for a long period (25-52 days), substantially later than their clinical recovery (12-37 days) (Supplementary Fig. 3). In this cohort, six children were older than 3 years of age, and should have relatively stable adult-like microbiome. The dynamic characteristics of the microbiome during COVID-19 implied that children’s microbiome is still particularly vulnerable and less resilient than that of the adults even after attaining a stable phase^7,9,16^. Importantly, the persistent worsening of the microbiomes caused by COVID-19 might impart potential short-term and long-term health problems during childhood and adulthood.

We and other have reported that altered respiratory microbiome with reduced bacterial diversity increased the susceptibility of children to acute respiratory tract infections (ARTIs)^6,8,25-27^. The impaired microbiome in children with COVID-19 is characterized by *Pseudomonas*-dominated community types that favor the colonization and growth of pathogenic and environmental bacteria, and a reduction of some beneficial commensals (Fig. 4). Probiotic (e.g. *Bifidobacterium*) and butyrate-producing bacteria (e.g. *Faecalibacterium*) that have anti-inflammatory ability were extensively depleted from the gut and respiratory microbiomes of the children especially at late stage of COVID-19 (Fig. 3b)^28,29^. Low abundance or lack of butyrate-producing bacteria might tolerate low-level inflammation induced by SARS-CoV-2, rendering children more vulnerable to ARTIs and diarrheal diseases^8,9,30^. In particular, the enrichment of genera *Moraxella* and *Streptococcus* in these COVID-19 children might predispose them to an increased risk of recurrent ARTIs^6^.

To the best of our knowledge, this is the first report on the complex dynamics of the gut and respiratory microbiota in children with COVID-19. Disturbed development of both gut and respiratory microbiomes, and prolonged respiratory dysbiosis caused by SARS-CoV-2 infection imply possible short-term and long-term complications after they have recovered from COVID-19, and predispose afflicted children to an increased health risk in later life. Although the long-term outcomes of COVID-19 on children need to be further studied with more extended follow-up and larger cohort, our data suggest that early implementation of various intervention strategies to modulate the microbiome development may provide clinical benefit to children in the post COVID-19 era.

### Online content

Any methods, additional references, source data, extended data, supplementary information, acknowledgements, details of author contributions and competing interests, and statements of data and number availability are available.

## Data Availability

Any methods, additional references, source data, extended data, supplementary information, acknowledgements, details of author contributions and competing interests, and statements of data and number availability are available. The raw data of 16S rRNA gene sequences are available at NCBI Sequence Read Archive (SRA) (https://www.ncbi.nlm.nih.gov/sra/) at BioProject ID PRJNA642019.

## Method

### Study population

Nine children were diagnosed as COVID-19 patients by Children’s Hospital of Fudan University according to the National Guidelines for Diagnosis and Treatment of COVID-19. A total of 103 samples, including 31 nasal swabs, 28 throat swabs and 44 feces, were collected from these patients (Supplementary Fig. S1). Twenty-five samples from 14 age-matched healthy children were used as controls. The respiratory samples were collected using flexible, sterile, dry swabs, which can reach the posterior nasopharynx and oropharynx easily (approximately 2 inches). About 2 ml spontaneous, unstimulated fecal specimen (300 mg/tube) was collected into a sterile cryogenic vial (Corning, NY, USA), divided into aliquots and stored at –80 °C until use. The sampling was performed by the professionals at the hospital.

The study was approved by Children’s Hospital of Fudan University (2020-27). Informed consents were obtained from involved patients or their guardians.

### Confirmation of COVID-19 children

The clinical and epidemiological characteristics, and SARS-CoV-2 RNA shedding patterns of these patients were previously reported^31,32^. All nine COVID-19 children had mild symptoms. The virus RNA was extracted from all samples using a Mag-Bind RNA Extraction Kit (MACCURA, Sichuan, China) according to the manufacturer’s instructions. The *ORFlab* and *N* genes of SARS-CoV-2 was detected using a Novel Coronavirus (2019-nCoV) RNA detection Kit (PCR-Fluorescence Probing) (DAAN gene, Guangzhou, China) according to the manufacturer’s instructions.

### 16S rRNA gene sequencing

All samples including nasal swabs, throat swabs and stool samples were subjected to the DNA extraction using a QIAamp DNA Microbiome Kit (QIAGEN, Düsseldorf, Germany). A novel triple-index amplicon sequencing strategy was used for 16S rRNA gene sequencing^33^. In brief, a set of universal bacterial primers was used to amplify the V4 hypervariable region (515-806 nt) of the 16S rRNA gene. Two rounds of PCR amplifications were performed ^34^. A reaction mixture containing 8 μL Nuclease-free water, 0.5 μL KOD-Plus-Neo (TOYOBO, Osaka Boseki, Japan), 2.5 μL of 1 μM PCR1 forward primer, 2.5 μL of 1 μM PCR1 reverse primer, and 5 μL DNA template was prepared for the first round of the PCR (PCR1) amplification. PCR1 products were purified using Monarch DNA Gel Extraction Kit (New England Biolabs, Ipswich, MA, USA), and quantified by a Qubit® 4.0 Fluorometer (Invitrogen, Carlsbad, CA, USA). Purified PCR1 products were pooled with equal amounts, and then subjected to the secondary round of PCR (PCR2) amplification. The PCR2 mix contains 21 μL Nuclease-free water, 1 μL KOD-Plus-Neo (TOYOBO, Osaka Boseki, Japan), 5 μL of 1 μM PCR2 forward primer, 5 μL of 1 μM PCR2 reverse primer, and 5 μL pooled PCR1 products. The PCR2 products were purified using the same Gel Extraction Kit and qualified using the Qubit® 4.0 Fluorometer. The specific products were further qualified using Agilent 2100 Bioanalyzer (Agilent, Santa Clara, CA, USA). The PCR2 products with equal moles of specific products were pooled and mixed them with AMPure XP beads (Beckman Coulter, Pasadena, CA, USA) in a ratio of 0.8:1. After purification, the amplicons were paired-end sequenced (2×250) using Illumina-P250 sequencer.

### Bioinformatic analysis of 16S rRNA gene sequence data

Using USEARCH11 software^35^ and FASTX-Toolkit^36^, sequenced reads were merged, de-multiplexed and filtered. After trimming barcode, adapter and primer sequences using USEARCH11, 14,702, 896 sequences were retained with an average of 100019 sequences per sample. Based on the Qiime2 platform^37^, the Deblur^38^ was used to cluster sequence data into sub-OTUs (operational taxonomic units), better than traditional OTU picking usually according to 97% sequence similarity threshold which may miss subtle and real biological sequence variation^39^. In particular, we used the Deblur to perform quality control, dereplicate, chimeras remove on with default settings except for truncating sequence length to 250bp. A sub-OTU (sOTU) count table, equivalent to OTU table, was generated (2187 sOTUs). The taxonomic classification of sOTU representative sequences was assigned by using the RDP Naive Bayesian Classifier algorithm^40^ based on the Ribosomal Database project (RDP) 16S rRNA training set (v16) database^41^. Finally, the sOTU table were subsampled at an even depth of 3600 sequences per sample to eliminate the bias led by different sequencing depths among samples. The sOTU coverage of 87% was sufficient to capture microbial diversity.

### Identification and characteristics of microbial community types

Based on the bacterial genus abundance, we used the Dirichlet multinomial mixtures (DMM)^42^ algorithm introduced by mothur^43^ v1.44.1 to identify microbial community types because the DMM algorithm could efficiently cluster samples based on microbial composition, whose sensitivity, reliability and accuracy were widely confirmed in many microbiome studies^16,44,45^. Based on the lowest Laplace approximation index, the appropriate microbial community type numbers (DMM clusters) were determined. Conjugated with the Analysis of Similarities (ANOSIM), the reliability of DMM clustering was further validated and then visualized by the Non-metric multidimensional scaling (NMDS) based on the Bray-Curtis distance under bacterial genus level. “The ANOSIM statistic “R” compares the mean of ranked dissimilarities between groups to the mean of ranked dissimilarities within groups. An R value close to “1.0” suggests dissimilarity between groups while an R value close to “0” suggests an even distribution of high and low ranks within and between groups^**46**^. ANOSIM p values lower than 0.05 suggest the higher similarity within sites. Richness (Observed sOTUs) and Pielou’s or Species evenness for each community type were calculated for estimating the difference of alpha diversity. Those analyses described above were performed using R package “vegan” v2.5-6. Using R package ‘Pheatmap’, the dynamic change of microbial community types and compositions were visualized. Alpha diversity difference between groups were measured using the Wilcoxon Rank Sum Test in R.

### Identification of indicator taxa contributing to microbial community typing

To get the reliable indicator genus for community typing, we performed the Indicator Species Analysis using the indicspecies package (ver.1.7.8) ^47^ in R platform with top 40 genus contributing to DMM clustering that accounted for 71% cumulative difference. Dynamic changes of indicator genera corresponding to each community type were showed in all COVID-19 children using the pheatmap package in R.

### Co-occurrence network analysis of microbiomes among gut, nasal, and throat of COVID-19 children

Based on microbial genus abundances of gut, nasal, and throat samples collected from 8 COVID-19 children, we calculated the Pearson Correlation Coefficient (Pearson’s r) among bacterial genera of three body sites. The Pearson’s r higher than 0.4 or lower than −0.4 with *P* values that were below 0.05 after the FDR adjustment were considered significant correlation. Co-occurrence network of significantly correlated bacterial genus pairs was visualized using Cytoscape v3.8.0^48^.

## Data availability

The raw data of 16S rRNA gene sequences are available at NCBI Sequence Read Archive (SRA) (https://www.ncbi.nlm.nih.gov/sra/) at BioProject ID PRJNA642019.

## Author contributions

C.Z. conceived the study idea. C.Z., Z.Z. and J.X designed and supervised the study. P.L., M.Z. and J.X collected clinical samples and data. R.X., P.L. and Y.M. performed the experiments. T.Z. and Q.W. processed and analyzed the raw sequencing data. R.X., P.L., J.X. and M.Z. analyzed the clinical data. C.Z., Z.Z. and X.J. interpreted the data. Z.Z. and R.X. generated the figures. C.Z., Z.Z., R.X., and T.Z. wrote the first draft of the manuscript. X.J. contributed to critical revision. All authors contributed to the final manuscript.

## Acknowledgements

We thank Mr. Kai Liu and Mrs. Xiuming Wu at Institut Pasteur of Shanghai, Chinese Academy of Sciences for their technical support. This work was supported by the grants from the National Key Research and Development Program of China (2017ZX10103009-002 and 2018YFC2000500), the Second Tibetan Plateau Scientific Expedition and Research (STEP) program (2019QZKK0503), the Key Research Program of the Chinese Academy of Sciences (FZDSW-219), the Chinese National Natural Science Foundation (31970571) and grants specific for Coronavirus Disease 2019 from the Children’s Hospital of Fudan University (Grant No. EKXGZX006).

## Additional information

**Supplementary Table S1. Fecal, nasal, and throat microbial abundances (phyla and genera)**. The stars represent unclassified genera.

**Supplementary Fig. S1. COIVD-19 patient admission and discharge times as well as the detection of SARS-CoV-2**. DAY 1 was the day of symptom onset.

**Supplementary Fig. S2. Dynamic change of 26 indicator genera in three body sites of nine COVID-19 children**. Linked to Fig.2a and Fig.3b. The stars represent unclassified genera.

**Supplementary Fig. S3. Distinct gut and respiratory microbiome mechanisms associated with the progress of COVID-19 in adults and children**. The dynamic mechanism of the microbiome in adults was interpreted from our recent observation based on the longitudinal throat and anal swabs from 35 adults with COVID-19 (Xu et al. unpublished data). Similar community types from I to III/IV indicate a progressive dysbiosis of the microbiome. In mild adults with COVID-19, a synchronous shift of community type from early dysbiosis towards late incomplete restoration was found in both respiratory and gut microbiomes within a short time. In children, however, the changes of the community types were divergent between the respiratory tract and gut, possibly implying that the “airway-gut axis” is still not established during the childhood. Moreover, children’s respiratory microbiome appeared to be progressively worsening for a long period despite a fast clinical recovery.

